# AKT1/mTOR/RICTOR risk variants in Indian hypertrophic cardiomyopathy patients

**DOI:** 10.64898/2026.07.23.26358782

**Authors:** Harshil Chittora, Dimple Notani, Perundurai S. Dhandapany

**Affiliations:** National Centre for Biological Sciences (NCBS), Tata Institute of Fundamental Research, Bengaluru, Karnataka, 560065, India; Cardiovascular Development and Disease Mechanisms, Institute for Stem Cell Science and Regenerative Medicine (BRIC-inStem), Bengaluru, Karnataka, 560065, India

## Abstract

Hypertrophic cardiomyopathy is a hereditary heart muscle disease characterized by abnormal ventricular thickening and is predominantly caused by mutations in sarcomeric and signaling genes. Despite these advances, a substantial proportion of patients carry no identifiable pathogenic variants in these genes. Recently, we have shown that mutations in the *RPS6KB1* gene (a member of the Akt signaling pathway) can lead to HCM. However, the genetic role of other members of the AKT pathway remains unknown in HCM. To address this gap, we used exome sequencing of an Indian-specific HCM patient cohort and identified six heterozygous missense variants in unrelated patients, including AKT1 (p.G37V), mTOR (p.A152S, p.D297N, p.R1818H), and RICTOR (p.R241Q, p.T1209M). The identified variants were either novel or ultra-rare and were classified as likely pathogenic according to ACMG guidelines. Functional consequences were evaluated using AKT1, mTOR, and RICTOR mutant proteins and compared with wild-type in a cardiomyocyte cell model. All six mutated proteins showed a significant increase in cell surface area, elevated mTOR signaling, induction of hypertrophic marker gene expression, and enhanced global protein synthesis, suggesting a gain-of-function effect. These findings underscore a potential genetic risk associated with the AKT/mTOR/RICTOR axis in patients with HCM.

## Introduction

Hypertrophic cardiomyopathy (HCM [MIM: 192600]) is a cardiac muscle disorder characterized by increased ventricular wall thickness and diastolic dysfunction (1,2). HCM is associated with mutations in genes encoding sarcomeric and signaling-associated proteins (3,4). However, genetic studies reveal the absence of reported cardiomyopathy-related pathogenic gene mutations in a significant number of patients (<50%) (3,5), suggesting that novel disease-associated genes remain to be discovered in the Indian population. Among the various hypertrophy-associated signaling pathways, the AKT/mTOR/RICTOR signaling axis is a key regulator of cardiac hypertrophy (6,7); yet pathogenic/likely pathogenic variants of uncertain significance (P/LP/VUS) in its core components have not been systematically investigated in the Indian HCM cohort. (5,6).

*AKT1* [MIM: 164730] encodes the AKT1 protein, a serine/threonine kinase that transmits signals from growth factors to phosphorylate and inhibit the TSC1/TSC2 complex [MIM: 605284]. This process removes the inhibition of Rheb GTPase [MIM: 601293], leading to the activation of mTOR (7,8). mTOR (*MTOR* [MIM: 601231]), in turn, drives protein synthesis through phosphorylation of ribosomal protein S6 kinase 1 (S6K1/p70S6K, encoded by *RPS6KB1* [MIM: 608938]), which regulates protein synthesis by phosphorylating the 40S ribosomal S6 protein (rpS6) (4,9), enabling the translational expansion necessary for cardiomyocyte hypertrophy(4,10). mTOR-interacting partners include other members, such as RAPTOR (*RPTOR* [MIM: 607130]) and RICTOR (*RICTOR* [MIM: 609022]), thereby enabling this process (11,12).

In this study, we describe six novel and ultra-rare heterozygous missense variants in AKT1 p.G37V (c.110G>T), mTOR p.A152S (c.454G>T), p.D297N (c.889G>A), and p.R1818H (c.5453G>A), and RICTOR p.R241Q (c.722G>A), p.T1209M (c.3626C>T) through whole-exome sequencing (WES) of a clinically well-characterized adult Indian HCM cohort (4,5). The identified variants were either absent or present at ultra-rare frequencies in the gnomAD South Asian control population (13). Our biochemical studies revealed that the newly identified variants induce cellular hypertrophy and pathological gene expression, global protein synthesis rate, and significantly activate rpS6 compared to wild-type conditions.

## Materials and methods

### Patient cohort

A total of 335 population-stratified, unrelated South Indian patients with primary HCM were used for this study. These well-characterized primary HCM patients were recruited from various tertiary hospitals, as previously described by us (5). Written consent was obtained from each of the subjects. Written consent was obtained from each of the subjects. The cases were diagnosed as HCM using a well-established methodology as proposed by the American Society of Echocardiography and Cardiac Magnetic Resonance Imaging (CMRI). Additional details, including the inclusion and exclusion criteria, are provided in our previous publications. (5,14). In brief, baseline characteristics of genotype-positive patients found in the cohort are detailed in Table 1.

**Table 1.** Detailed characteristics of the genotype-positive Indian HCM patients.

| Clinical Parameters | AKT1<br>p.G37V | mTOR<br>p.A152S | mTOR<br>p.D297N | mTOR<br>p.R1818H | RICTOR<br>p.R241Q | RICTOR<br>p.T1209M |
| --- | --- | --- | --- | --- | --- | --- |
| Age | Adult | Adult | Adult | Adult | Adult | Adult |
| HTN | No | No | No | No | No | No |
| Heart disease (HCM) | Yes | Yes | Yes | Yes | Yes | Yes |
| LVEF% | 65 | 68 | 72 | 69 | 65 | 70 |
| IVSd, mm | 19 | 18 | 21 | 22 | 20 | 22 |
| IVSs, mm | 23 | 24 | 25 | 23 | 22 | 21 |
| CAD | No | No | No | No | No | No |
| Cancer | No | No | No | No | No | No |
| Syndrome | No | No | No | No | No | No |
| Sarcomeric variants | - | - | - | MYH7<br>p.R453C | MYBPC3<br>p.C1124X | - |

### Whole-exome and targeted sequencing

Genomic DNA was recovered from peripheral blood lymphocytes or mononuclear cells using standard extraction procedures. Sequencing libraries were constructed and enriched for the target regions with the SureSelect V5 Target Enrichment kit (Agilent Technologies), then amplified by PCR. Sequencing was performed by external service providers and on the in-house sequencing facility, with patient identifiers removed beforehand. Each exome was sequenced to generate paired-end 100-base-pair reads at approximately 100× coverage.

Low-quality reads were discarded, and adapter sequences were trimmed with Trimmomatic (v0.32) (15). Cleaned reads were aligned to the human reference genome (GRCh38) using the Burrows–Wheeler Aligner (v0.7)(16). PCR duplicates were removed with Picard (v2.9.0). Variants were called with the HaplotypeCaller module of the Genome Analysis Toolkit (17) (version 4.1.9.0; Broad Institute, Cambridge, MA) and annotated using the ANNOVAR framework (18).

Candidate gene variants were filtered based on predicted pathogenicity using an ensemble of in silico predictors, including PolyPhen-2, SIFT, PANTHER-PSEP, MetaDome, CADD, MutationTaster, and M-CAP. (19–23). A variant was considered damaging when at least four of seven predictors flagged it. Prioritization was given to rare coding variants (minor allele frequency ≤ 0.1%), ultra-rare (≤ 0.01%), or entirely novel across public reference datasets spanning multiple ancestries (gnomAD global). Only novel and rare variants were retained, and final classification as pathogenic, likely pathogenic, or variant of uncertain significance (VUS) followed ClinVar and ACMG guidance. (24). Candidate variants were confirmed by amplifying the relevant exons and flanking intronic boundaries from genomic DNA. PCR products were purified using a column-based kit (QIAGEN), Sanger-sequenced, and confirmed.

### Expression constructs

The following expression constructs were obtained from Addgene: pEGFP-Akt1(WT) (plasmid #86637; http://n2t.net/addgene:86637; RRID: Addgene_86637), which was a gift from Thomas Leonard and Ivan Yudushkin(25); pRK5-HA-YFP-rictor (plasmid #73387; http://n2t.net/addgene:73387; RRID:Addgene_73387), which was a gift from Jie Chen and Taekjip Ha(26); and pcDNA3-Flag mTOR wt (plasmid #26603; http://n2t.net/addgene:26603; RRID:Addgene_26603), which was a gift from Jie Chen(27). Variant plasmids for AKT1, mTOR, and RICTOR were generated by site-directed mutagenesis from sequence-verified parental constructs. All mutations were confirmed by Sanger sequencing. Tagged protein expression was verified by immunoblot against GFP (AKT1), FLAG (mTOR), and HA (RICTOR).

### Cell culture and transfection

H9c2 rat cardiomyocytes (ATCC CRL-1446) were cultured in DMEM supplemented with 10% FBS at 37°C, 5% CO₂. Cells were transfected at 50-60% confluency using jetPrime transfection reagent (Sartorius, #101000001) for 48 hours per manufacturer’s instructions.

### Immunofluorescence and cell surface area measurement

Cells were fixed (4% paraformaldehyde, 15 min, room temperature), permeabilized (0.1% Triton X-100, 10 min), blocked (5% BSA, 1 h), and stained with Phalloidin 488 (1:200; Thermo Fisher Scientific, #A12379) for 1 hour at room temperature. Nuclear staining was performed using Hoechst dye (Thermo Fisher Scientific, #62249). After washing cells twice, coverslips were mounted using VECTASHIELD antifade mounting medium (Vector Laboratories, #H-1000). Images were collected using the Olympus IX73 inverted fluorescence microscope system and analyzed in ImageJ (Fiji) from≥30-50 cells per condition across three independent experiments.

### Immunoblotting

Cells were lysed in RIPA buffer supplemented with protease and phosphatase inhibitor cocktails (Thermo Fisher Scientific, #A32961). Protein concentration was determined by BCA assay (Thermo Fisher Scientific, #23225), and equal amounts (20–30 µg) were resolved by SDS-PAGE and transferred to PVDF membranes (Biorad, #1620177). Membranes were blocked in 5% BSA/TBS-T and probed overnight at 4 °C with the following primary antibodies (all 1:1,000): phospho-rpS6 (Ser240/244) (clone G.22.6, rabbit monoclonal; Thermo Fisher Scientific, #701845); total rpS6 (clone 54D2, mouse monoclonal; Cell Signaling Technology [CST], #2317); phospho-mTOR (Ser2448) (clone D9C2, rabbit monoclonal; CST #5536); total mTOR (clone 7C10, rabbit monoclonal; CST #2983); phospho-Akt (Ser473) (clone D9E, rabbit monoclonal; CST #4060); total Akt (clone C67E7, rabbit monoclonal; CST #4691); GFP (clone D5.1, rabbit monoclonal; CST #2956); DYKDDDDK (FLAG) Tag (rabbit polyclonal; CST #2368); HA-Tag (clone 2-2.2.14, mouse monoclonal; Thermo Fisher Scientific, #26183); and GAPDH (loading control; mouse monoclonal; Thermo Fisher Scientific, #MA5-15738). After washing, membranes were incubated with HRP-conjugated goat anti-rabbit (1:10,000; Thermo Fisher Scientific, #31460) and goat anti-mouse IgG (1:10,000; Thermo Fisher Scientific, #31430) secondary antibodies, and signals were visualized by enhanced chemiluminescence (Clarity Western ECL Substrate, Biorad, #1705061). Phospho-blots were stripped and re-probed for the corresponding total protein to confirm equal loading, with GAPDH as the internal loading control. Bands were quantified by densitometry in ImageJ (NIH), normalizing the phosphoprotein signal to total protein in the same lane; data are expressed as fold change relative to wild type.

### Quantitative RT-PCR

Total RNA was extracted with TRIzol (Thermo Fisher Scientific, #15596026) and reverse-transcribed using Verso cDNA synthesis kit (Thermo Fisher Scientific, #AB1453A). Hypertrophic marker expression *Nppa*, *Nppb*, *Myh6,* and *Myh7* was quantified by qRT-PCR using PowerUp™ SYBR™ Green Master Mix (Thermo Fisher Scientific, # A25742). Data were normalized to *ACTB* by the 2^⁻ΔΔCt^ method and expressed as fold-change relative to wild-type. Reactions were performed in triplicate from three independent experiments.

### SUnSET global protein synthesis assay

Global protein synthesis was quantified by surface sensing of translation (SUnSET) assay. After 48 hours of transfection, the medium was aspirated, and cells were washed twice with pre-warmed sterile 1× PBS. A puromycin premix was prepared in pre-warmed medium and added to the cells at a final concentration of 1 µM for 30 minutes under normal culture conditions. Following the pulse, the puromycin-containing medium was aspirated, and cells were washed three times with ice-cold 1× PBS. Plates were placed on ice, and cells were scraped into RIPA buffer for protein isolation.

Equal protein amounts (at least 20–30 µg per lane) were normalized to equal volumes with gel running buffer, combined with an equal volume of 2× Laemmli sample buffer containing 5% (v/v) β-mercaptoethanol, and boiled at 95 °C for 5 minutes. Puromycin-labeled nascent polypeptides were detected by anti-puromycin immunoblot (clone 12D10; Merck KGaA, Darmstadt, Germany). Total protein (Ponceau S) served as loading normalization. A no-puromycin lane was included in each experiment as a specificity control.

### Statistical analysis

Continuous baseline characteristics are reported as mean ± standard deviation. Group comparisons in the functional experiments used a two-tailed Student’s t-test or a one-way analysis of variance with the appropriate post hoc test (Dunnett’s), and each experiment was performed in triplicate. Categorical comparisons of variant burden, including case-excess and odds-ratio estimates with 95% confidence intervals, were evaluated by Fisher’s exact test. Multiple-comparison adjustment was performed using the Benjamini–Hochberg procedure, and statistical significance was set at an adjusted P-value below 0.05.

## Results

### AKT1/mTOR/RICTOR variants associated with HCM

We analyzed whole-exome sequencing data from 335 well-characterized Indian adult HCM probands using our in-house variant-filtering pipeline(4,5), with a focus on members of the mTOR pathway. The details of the patient’s clinical features are given in Table 1. We identified six heterozygous missense variants in three different genes encoding *AKT1, MTOR*, and *RICTOR*. We didn’t observe any potential P/LP/VUS variants in other pathway members, including *RPTOR* [MIM: 607130], *DEPTOR* [MIM: 612974], *MLST8* [MIM: 608221], *MAPKAP1* [MIM: 610853], and *PRR5* [MIM: 610121]. Additional HCM-related variants present in genotype-positive patients are given in Table 1. The AKT1 variant p.G37V was identified in one proband (Figure 1A). This variant was absent in the gnomAD control set, including South Asians (SA), and was predicted to be pathogenic by various *in silico* tools (Table 2). Three mTOR variants were identified in three different patients: p.A152S, p.D297N, and p.R1818H. The mTOR p.A152S was novel, and the other two variants were present in gnomAD SA had MAF < 0.01%. Two RICTOR variants were identified in two different patients, with p.R241Q absent from gnomAD SA and p.T1209M with gnomAD SA MAF < 0.01%. All variants were validated by Sanger sequencing, which confirmed their heterozygous state in affected probands (Figure 1A).

**Figure 1.**
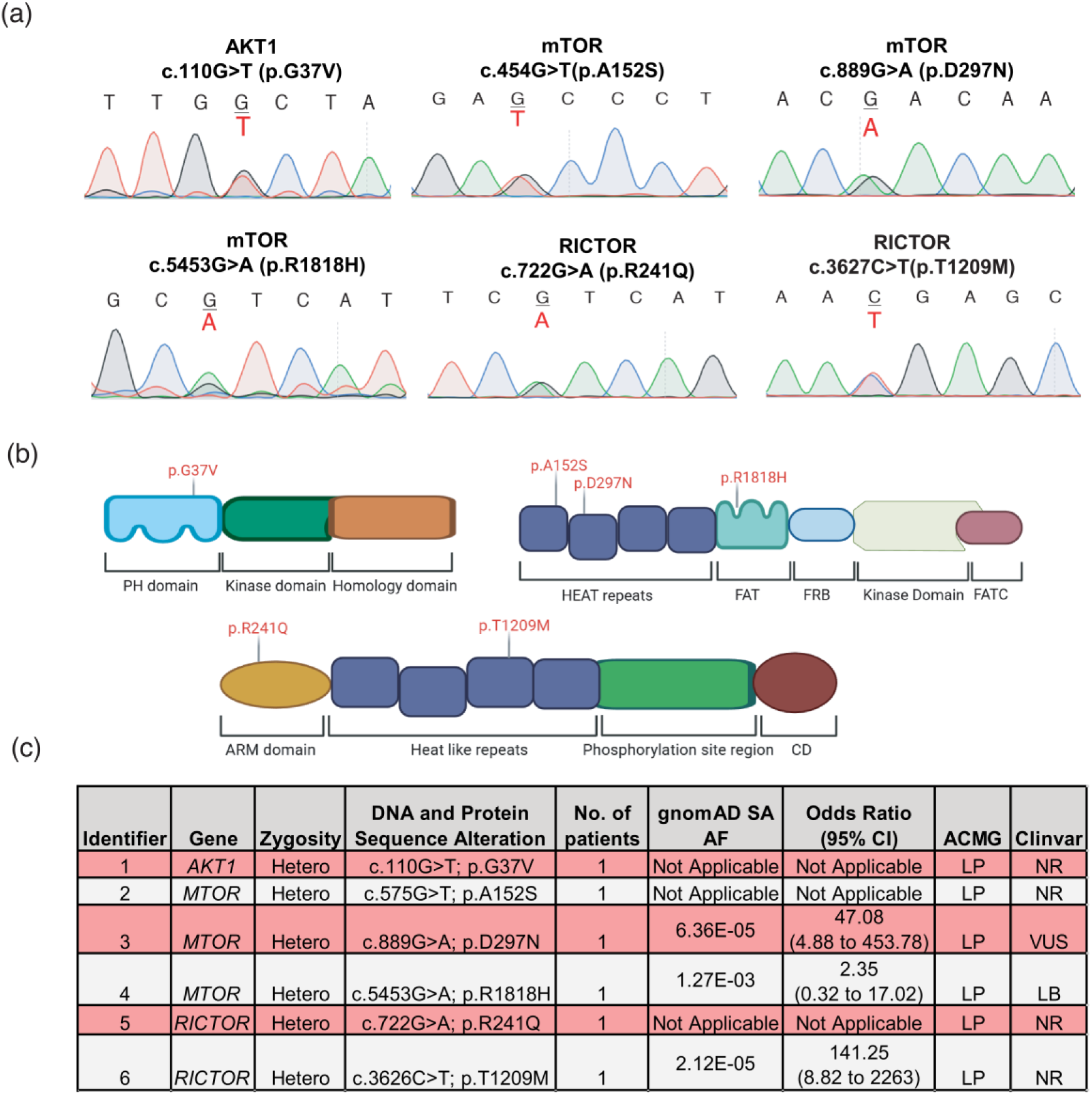
Identification of rare heterozygous variants in AKT1, MTOR, and RICTOR in patients with cardiomyopathy. **(a)** Representative Sanger sequencing chromatograms confirming the six heterozygous variants identified by exome sequencing: AKT1 c.110G>T (p.G37V), mTOR c.454G>T (p.A152S), mTOR c.889G>A (p.D297N), mTOR c.5453G>A (p.R1818H), RICTOR c.722G>A (p.R241Q) and RICTOR c.3627C>T (p.T1209M). The altered nucleotide is indicated in red beneath the reference base at each position. **(b)** Schematic representation of the AKT1 (top left), mTOR (top right), and RICTOR (bottom) protein domain architectures showing the position of each variant (red). **(c)** Summary of the identified AKT1, mTOR and RICTOR variants. (gnomAD SA – gnomAD South Asian)

**Table 2.** In silico analysis of the *AKT*, *MTOR,* and *RICTOR* gene variants using various computational tools.

| *in silico* prediction tool | AKT1<br>p.G37V | mTOR<br>p.A152S | mTOR<br>p.D297N | mTOR<br>p.R1818H | RICTOR<br>p.R241Q | RICTOR<br>p.T1209M |
| --- | --- | --- | --- | --- | --- | --- |
| SIFT_pred | D | D | T | D | D | D |
| Polyphen2_HDIV_pred | D | P | D | D | D | P |
| MutationTaster_pred | D | D | D | D | D | D |
| CADD_phred | 31 | 24 | 25.7 | 28.5 | 31 | 27.5 |
| M-CAP_pred | D | D | D | D | D | T |
| PANTHER-PSEP | D | D | T | T | D | D |
| Metadome | D | D | D | D | D | D |

(D damaging, P possibly damaging, T tolerated)

Protein domain mapping of the variants revealed that AKT1 p.G37V resides within the N-terminal Pleckstrin Homology (PH) domain, which is essential for PI3K-generated PIP3-dependent membrane recruitment and AKT1 activation (25,28). The three mTOR variants localize to the HEAT repeat domain (p.A152S, p.D297N) and the FAT domain (p.R1818H), regions critical for protein–protein interactions within the mTOR complexes (29,30). The two RICTOR variants, p.R241Q and p.T1209M, reside within the Armadillo (ARM) repeat-containing domain and the C-terminal PR domain, respectively, both of which are essential for mTOR complex assembly and substrate binding (31,32) (Figure 1B). *In silico* prediction algorithms classified these variants as damaging across multiple tools; CADD PHRED scores ranged from 24 to 31, all exceeding the conventional threshold of 20 (Table 2). Two patients carried concurrent sarcomere variants; both are noted in Table 1.

### AKT1/mTOR/RICTOR variants induce cellular hypertrophy

To understand the functional consequences of the identified variants, we expressed WT and mutant AKT1, mTOR, and RICTOR plasmids in the H9c2 cardiomyocyte cell line. Expression of AKT1 p.G37V significantly increased cell surface area relative to AKT1 WT expressing cells (Figure 2a). Among the mTOR variants, all three p.A152S, p.D297N, and p.R1818H significantly enlarged cell surface area relative to mTOR WT (Figure 2b). RICTOR p.R241Q and RICTOR p.T1209M variants significantly increased cell surface area relative to RICTOR WT (Figure 2c).

**Figure 2.**
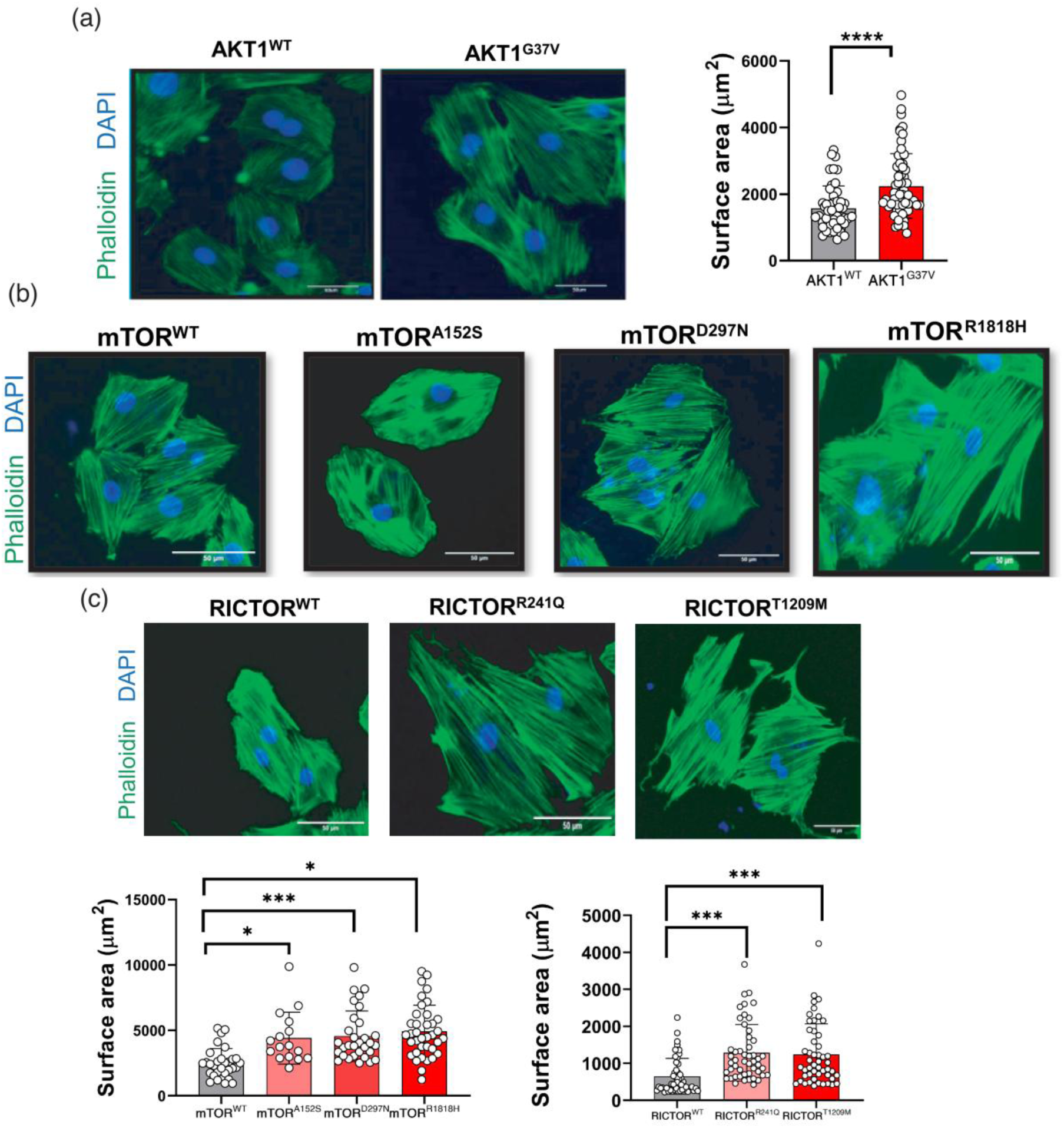
AKT1, mTOR, and RICTOR variants induce cellular hypertrophy in vitro. Representative immunofluorescence images of cells expressing wild-type or mutant constructs, stained with phalloidin to visualize F-actin (green) and DAPI to label nuclei (blue). Scale bars, 50 µm. **(a)** Cells expressing AKT1 WT or AKT1 p.G37V, with quantification of cell surface area (right). **(b)** Cells expressing mTOR WT, mTOR p.A152S, mTOR p.D297N, or mTOR p.R1818H. **(c)** Cells expressing RICTOR WT, RICTOR p.R241Q, or RICTOR p.T1209M. Lower panels show quantification of cell surface area (µm²) for the mTOR (left) and RICTOR (right). Error bars indicate mean ± SEM from n=3 (50 cells each) independent experiments. Statistical significance was determined by Student’s unpaired t-test or one-way ANOVA with Dunnett’s post hoc test versus the corresponding wild-type control. *P < 0.05; ***P < 0.001; ****P < 0.0001.

### AKT1/mTOR/RICTOR variants increase protein synthesis rate

AKT1/mTOR/RICTOR activates downstream phosphorylation of rpS6, resulting in global protein synthesis(4,9). In the hypertrophic heart, this is a critical mediator of the increase in cardiomyocyte size that leads to heart wall thickening (10,33). To determine the protein synthesis rate, we employed the SUnSET (Surface Sensing of Translation) assay, in which puromycin is incorporated into nascent polypeptide chains as a quantitative proxy for the global translation rate (34). Immunoblot analysis of cardiomyocyte lysates expressing AKT1, mTOR, or RICTOR mutants showed significant increases in protein synthesis across all six variants compared with their respective wild-type controls (Figure 3). AKT1 p.G37V increased puromycin incorporation relative to AKT1 WT (Figure 3a). All three mTOR variants produced significant increases (Figure 3b). Both RICTOR variants similarly enhanced puromycin incorporation (Figure 3c).

**Figure 3.**
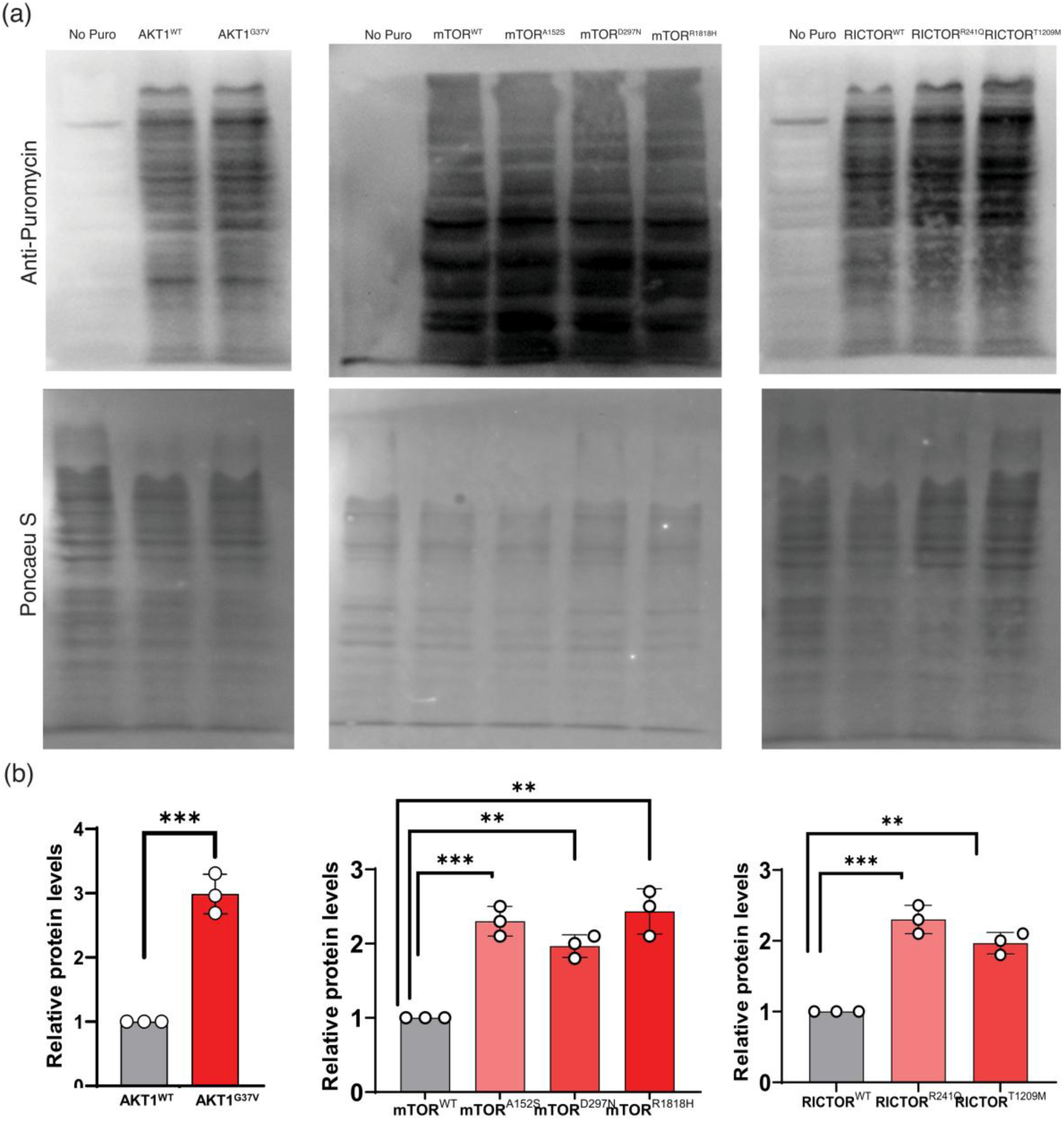
Expression of AKT1, mTOR, and RICTOR variants increases global protein synthesis. **(a)** Representative SUnSET assays in cells expressing wild-type or mutant AKT1 (left), mTOR (middle), or RICTOR (right). Newly synthesized proteins were labeled with puromycin and detected by immunoblotting with an anti-puromycin antibody (upper panels). Puromycin-untreated cells (No Puro) served as a negative control. Ponceau S staining of the membrane (lower panels) is shown as a loading control. **(b)** Densitometric quantification of total puromycin incorporation, normalized to Ponceau S signal and expressed relative to the corresponding wild-type control. Bars indicate mean ± SEM of three independent experiments; individual data points are shown. Statistical significance was determined by Student’s unpaired t-test / one-way ANOVA with Dunnett’s post hoc test versus wild type. **P < 0.01; ***P < 0.001.

### Elevated fetal gene and rpS6 phosphorylation by AKT1/mTOR/RICTOR variants

We next examined the reactivation of the embryonic gene program and the activation status along the AKT1/mTOR cascade. Quantitative RT-PCR of hypertrophic marker genes, presented as expression heatmaps, showed induction of the fetal program across all three genotypes relative to wild-type (Figure 4a). AKT1 p.G37V induced *NPPA* [MIM: 108780], *NPPB* [MIM: 600295], *MYH6* [MIM: 160710], and *MYH7* [MIM: 160760]. All the mTOR variants elicited the induction of hypertrophy markers, consistent with the impact on the morphology and protein synthesis. Both RICTOR variants elevated *NPPA*, *NPPB*, *MYH6,* and *MYH7* relative to wild-type.

**Figure 4.**
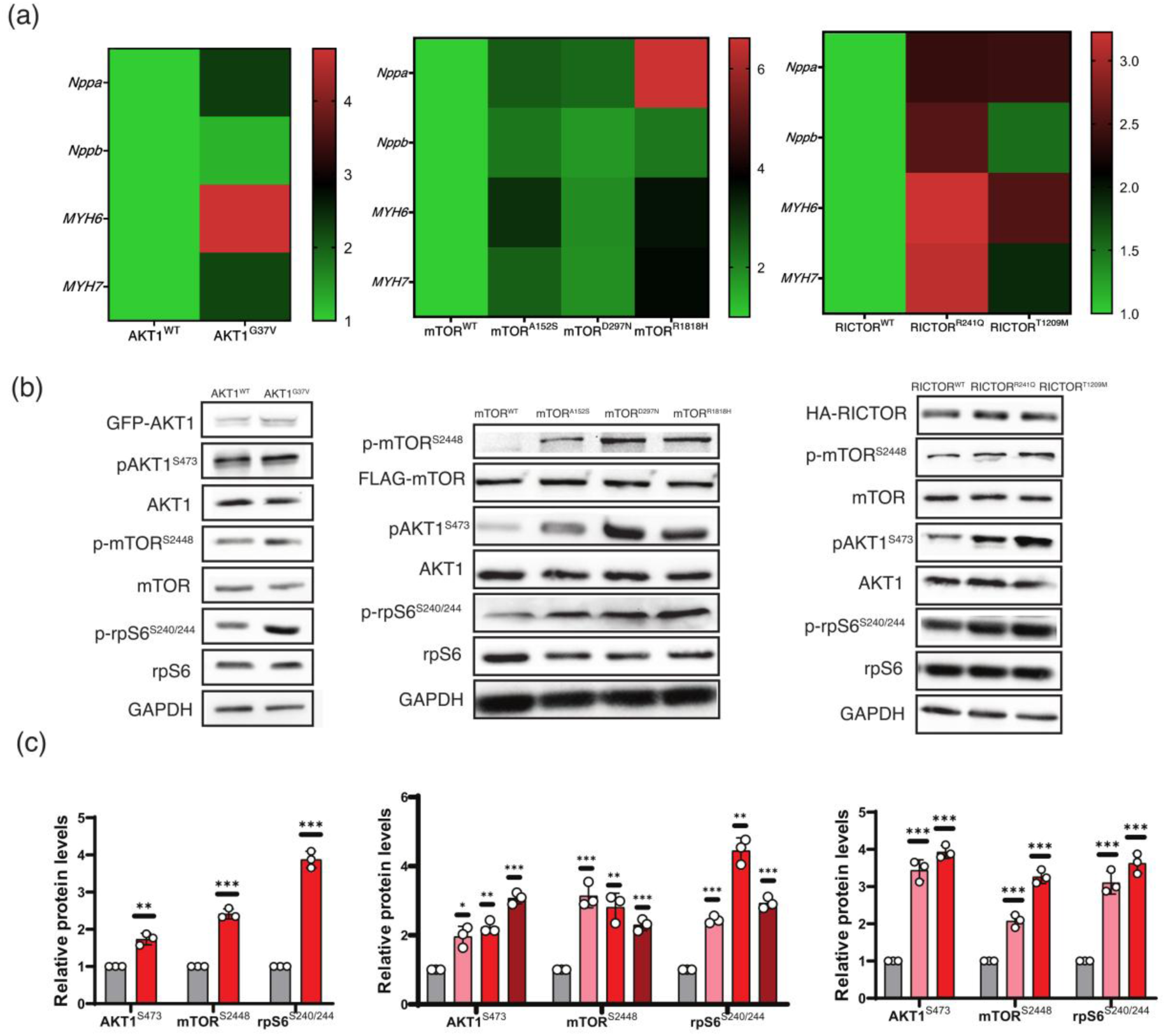
Variants activate AKT–mTOR signalling and induce a hypertrophic gene expression program. **(a)** Heatmaps showing relative mRNA expression of hypertrophic marker genes in cells expressing wild-type or mutant AKT1 (left), mTOR (middle), or RICTOR (right), quantified by qRT-PCR and normalized to *ACTB* and to the corresponding wild-type control. Color scale indicates fold change relative to wild type. ANP, atrial natriuretic peptide (*NPPA*); BNP, brain natriuretic peptide (*NPPB*); α-MHC, α-myosin heavy chain (*MYH6*); β-MHC, β-myosin heavy chain (*MYH7*) **(b)** Representative immunoblots of AKT/mTOR pathway components in cells expressing wild-type or mutant AKT1 (left), mTOR (middle), or RICTOR (right). GAPDH served as a loading control. **(c)** Densitometric quantification of phosphorylated AKT1 (Ser473), mTOR (Ser2448), and rpS6 (Ser240/244), each normalized to the corresponding total protein and expressed relative to wild type. Bars indicate mean ± SEM of three independent experiments; individual data points are shown. Statistical significance was determined by a post hoc test versus the wild type. *P < 0.05; **P < 0.01; ***P < 0.001.

To understand the signaling mechanisms, we assessed phospho-AKT1 (Ser473), phospho-mTOR (Ser2448), and phospho-rpS6 (Ser240/244) (Figure 4b) as needed. Robust expression of each construct was confirmed with the respective tag antibodies (GFP for AKT1, FLAG for mTOR, HA for RICTOR), and rpS6 phosphorylation was assessed in parallel; GAPDH served as the loading control.

Densitometric quantification (Figure 4c) revealed that all six variants increased rpS6 phosphorylation compared to their wild-type controls. AKT1 p.G37V-expressing cells showed elevated phospho-AKT1 (Ser473), phospho-mTOR (Ser2448) and phospho-rpS6 (Ser240/244). The three mTOR variants produced graded increases across the same three proteins. Both RICTOR variants (R241Q and T1209M) likewise elevated phospho-AKT1 (Ser473), phospho-mTOR (Ser2448), and phospho-rpS6 (Ser240/244). Notably, Ser473 in AKT1 is a RICTOR-sensing phosphorylation site that promotes hypertrophy.

## Discussion

The AKT/mTOR signaling cascade is critical for cardiomyocyte growth. (10,33). While various studies have demonstrated the involvement of this pathway in cardiac hypertrophy, there is a lack of research examining genetic perturbations in its members associated with HCM. In this study, we performed genetic screening of clinically well-characterized Indian primary HCM probands, identified six rare heterozygous missense variants distributed across three genes encoding core components of the AKT/mTOR/RICTOR signaling axis: AKT1 (p.G37V), mTOR (p.A152S, p.D297N, p.R1818H), and RICTOR (p.R241Q, p.T1209M). These variants account for 1.8% of Indian HCM patients (6 out of a total of 335 patients). All six variants were either absent or present at ultra-rare frequency in the gnomAD v4.1 South Asian population and were classified as likely pathogenic by the ACMG/AMP criteria. (35). Three of the six variants were novel, and the remaining three demonstrated case enrichment compared with gnomAD South Asian controls, including mTOR (p.D297N: OR=47.08; 95% CI 4.88 to 453.78; p=0.0009 and p.R1818H: OR= 2.35; 95% CI 0.32 to 17.02; p=0.3972) and RICTOR (p.T1209M: OR=141.25; 95% CI 8.82 to 2263; p=0.0005) compared to gnomAD South Asian controls (Figure 1C).

No pathogenic or likely pathogenic variants were identified in other mTOR pathway members, including *RPTOR, DEPTOR, MLST8, MAPKAP1,* and *PRR5*. Notably, none of the six variant carriers exhibited other clinical features of known *AKT1*, *MTOR*, or *RICTOR*-associated syndromes, distinguishing the phenotype observed in our cohort. This might be due to positional effects of the variants and interaction between other modifier genes or environmental factors (1,36). However, our functional data suggest that the variants can independently induce cardiac hypertrophy by increasing cell size, activating the fetal gene program, and increasing mTOR signaling.

The SUnSET assay data provide direct evidence that all six variants elevate global protein synthesis, a critical and rate-limiting determinant of cardiomyocyte mass accumulation in pathological hypertrophy(10,34). The approximately threefold increase in puromycin incorporation in AKT1 p.G37V cells, and the significant increases observed across all mTOR and RICTOR variants, are consistent with mTOR-driven stimulation of ribosomal biogenesis and cap-dependent mRNA translation. Sustained elevation of protein synthesis, rather than the transient increase associated with physiological hypertrophy, is a hallmark of the pathological hypertrophic response, and the overall enhancement observed across all six variants supports a shared translational mechanism underlying the convergent morphological phenotype.

The AKT1/mTOR axis is a canonical driver of cell growth, and activating variants in each of these genes are already recognized in developmental overgrowth and neurodevelopmental disorders, but through variants that differ markedly from those we report. AKT1, recruited to the membrane via its PH domain following PI3K-generated PIP3 signaling, phosphorylates and inactivates the TSC1/TSC2 complex to relieve inhibition of Rheb and activate mTOR. The pathogenic disease allele of Proteus syndrome [MIM: 176920], p.E17K in the AKT1 PH domain, locks AKT1 at the membrane and constitutively activates it, leading to asymmetric somatic overgrowth, epidermal naevi, and vascular malformations(37). However, the HCM-associated p.G37V genotype-positive patient didn’t show any signs of Proteus syndrome. This might be due to differential binding affinity to the cell membrane, as reported earlier (38). Strongly activating MTOR variants that majorly cluster in the catalytic (kinase) domain are associated with Smith–Kingsmore syndrome [MIM: 616638](39,40). HCM-related MTOR variants, p.A152S and p.D297N, fall in the N-terminal HEAT repeats and p.R1818H in the FAT domain, which is different from Smith–Kingsmore syndrome hotspots.

RICTOR, the defining scaffold of mTORC2 that phosphorylates AKT at Ser473 and coordinates substrate interactions across the network, has been tentatively linked to intellectual disability and neurodevelopmental delay via mTORC2 dysregulation(31,41), with no previously described cardiac phenotype. In contrast, HCM-related RICTOR variants, p.R241Q and p.T1209M, occupy the ARM repeats and C-terminal region domains that mediate complex assembly and substrate scaffolding rather than direct catalysis (12,32).

Our functional data are consistent with a substantial body of literature implicating the AKT– mTOR pathway in pathological cardiac hypertrophy across different model systems. (33,42,43). Cardiac-specific overexpression of constitutively active AKT in mice produces concentric hypertrophy that progresses to heart failure(42), while cardiac-specific deletion of AKT1 attenuates pressure-overload-induced hypertrophy (44). Cardiac-specific mTOR activation via TSC2 deletion drives pathological hypertrophy and fibrosis (8,44), and rapamycin treatment reverses established hypertrophy across multiple rodent models (45). The elevation of phospho-rpS6 (Ser240/244) across all six variants is particularly informative, as rpS6 phosphorylation by S6K1 is a prominent readout of mTOR complex activity, and its sustained elevation was previously documented by other groups and us(4,45). Notably, we recently reported that variants in RPS6KB1 hyperactivate rpS6, leading to HCM (4).

Two patients in our cohort carried concurrent sarcomere variants, MTOR p.R1818H co-occurring with MYH7 p.R453C, and RICTOR p.R241Q co-occurring with MYBPC3 p.C1124X, which precludes exclusive attribution of HCM to the AKT/mTOR variants in those individuals. However, the independent functional demonstration that each AKT/mTOR variant drives hypertrophy in a controlled cellular system supports intrinsic pathogenicity regardless of the concurrent sarcomere variant(4). These cases may represent a digenic architecture, at least in a subset of patients, in which both variants act additively to modulate penetrance and phenotypic severity, consistent with accumulating evidence that HCM expressivity is influenced by multiple variant burden across many loci and that modifier variants in signaling genes may amplify sarcomere-driven hypertrophy(36,46).

In conclusion, we demonstrated that variants across *AKT1, MTOR,* and *RICTOR* identified in clinically well-defined, non-syndromic primary Indian HCM patients produce a cardiomyocyte hypertrophy phenotype characterized by enhanced global protein synthesis, activation of fetal gene reprogramming, and activation of the mTOR pathway. These findings emphasize the need to analyze the AKT/mTOR/RICTOR pathway variants and their role in HCM patients.

## Data Availability Statement

Data are available from the corresponding author upon reasonable request. All data relevant to the study are included in the article.

## Acknowledgments

The authors thank Addgene (Plasmid #86637, Plasmid #73387, and Plasmid #26603).

## Author Contribution Statement

PSD conceived, designed, analyzed, and drafted the manuscript. HC performed, designed, and analyzed the major functional studies. PSD and DN provided reagents for the study.

## Ethical Approval

This study was approved by the Institutional Ethical Committee of the Institute for Stem Cell Science and Regenerative Medicine, Bangalore (Reference number: inStem/IEC-10/001). Informed consent for the use of clinical data, blood, and/or tissue samples for the study was obtained from the participants. The institutional review boards of the study centers approved the study protocols.

